# Impact of transseptal puncture location on the fossa ovalis on first-pass pulmonary vein isolation

**DOI:** 10.1101/2024.07.18.24310668

**Authors:** Kohei Matsunaga, Tadashi Hoshiyama, Shozo Kaneko, Hitoshi Sumi, Hisanori Kanazawa, Yuta Tsurusaki, Yuichiro Tsuruta, Masanobu Ishii, Shinsuke Hanatani, Hiroki Usuku, Eiichiro Yamamoto, Yasuhiro Izumiya, Kenichi Tsujita

## Abstract

**Background:** Recently, radiofrequency catheter ablation (RFCA) has become an important treatment strategy for atrial fibrillation (AF). During this procedure, achieving first-pass pulmonary vein (PV) isolation–PV isolation in which no residual conduction gap remains following initial circumferential lesion is created around the PV–has proven to lead better results in terms of AF recurrence. Although various risk factors for the creation of residual conduction gap have been proposed, the relationship between the transseptal puncture location on fossa ovalis and first-pass PV isolation success rate has not been clarified. Therefore, we investigate the relationship through this investigation.

**Methods:** Overall, 102 consecutive patients who had undergone their first RFCA for AF were included. These patients were divided based on the transseptal puncture location (infero-anterior, infero-posterior, supero-anterior, and supero-posterior), which was confirmed by imaging of three-dimensional structure of the anatomical fossa ovalis creating intracardiac echocardiography. The relationship between transseptal puncture location and the first-pass PV isolation success rate was analyzed.

**Results:** Among all 102 patients, number of transseptal puncture location were located in infero-anterior, infero-posterior, supero-anterior, and supero-posterior were 26, 61, 6, and 9 respectively. Among these, first-pass PV isolation success rate in the infero-posterior group exhibited the highest 79% (48/61 patients) compared to that in other locations [infero-anterior 61% (16/26 patients), supero-anterior 33% (2/6 patients), and supero-posterior 44% (4/9 patients); P=0.02]. Regarding ablation parameters, although the ablation index was not significantly different between each group (infero-anterior 401.6±7.6, infero-posterior 401.9±5.2, supero-anterior 397.5±4.7, and supero-posterior 398.6±5.3; P = 0.176). The P-vector, which represents insufficient catheter contact, was significantly observed lower frequency in the infero-posterior group (8.6%; P < 0.01) than in the other groups.

**Conclusion:** The transseptal puncture location in PV isolation is an important factor to achieve first-pass PV isolation, and it might affect AF recurrence.

**Non-standard Abbreviations and Acronyms:** AF, atrial fibrillation; AI, ablation index; FAM, fast anatomical map; PV, pulmonary vein; RFCA, radiofrequency catheter ablation

Clinical Perspective

**What is Known?:**

- Although transseptal puncture is an essential technique in atrial fibrillation ablation, the relationship between the transseptal puncture location on the fossa ovalis and first-pass pulmonary vein isolation success rate has not been fully evaluated so far.

**What the Study Adds:**

- Transseptal puncture at the infero-posterior region on the fossa ovalis was shown to result in a higher first-pass PV isolation proportion than that in other locations, owing to the better catheter contact situation.
- When performing catheter ablation for atrial fibrillation, it should be kept in mind that the transseptal puncture location might affect atrial fibrillation recurrence.

**Graphic abstract:** 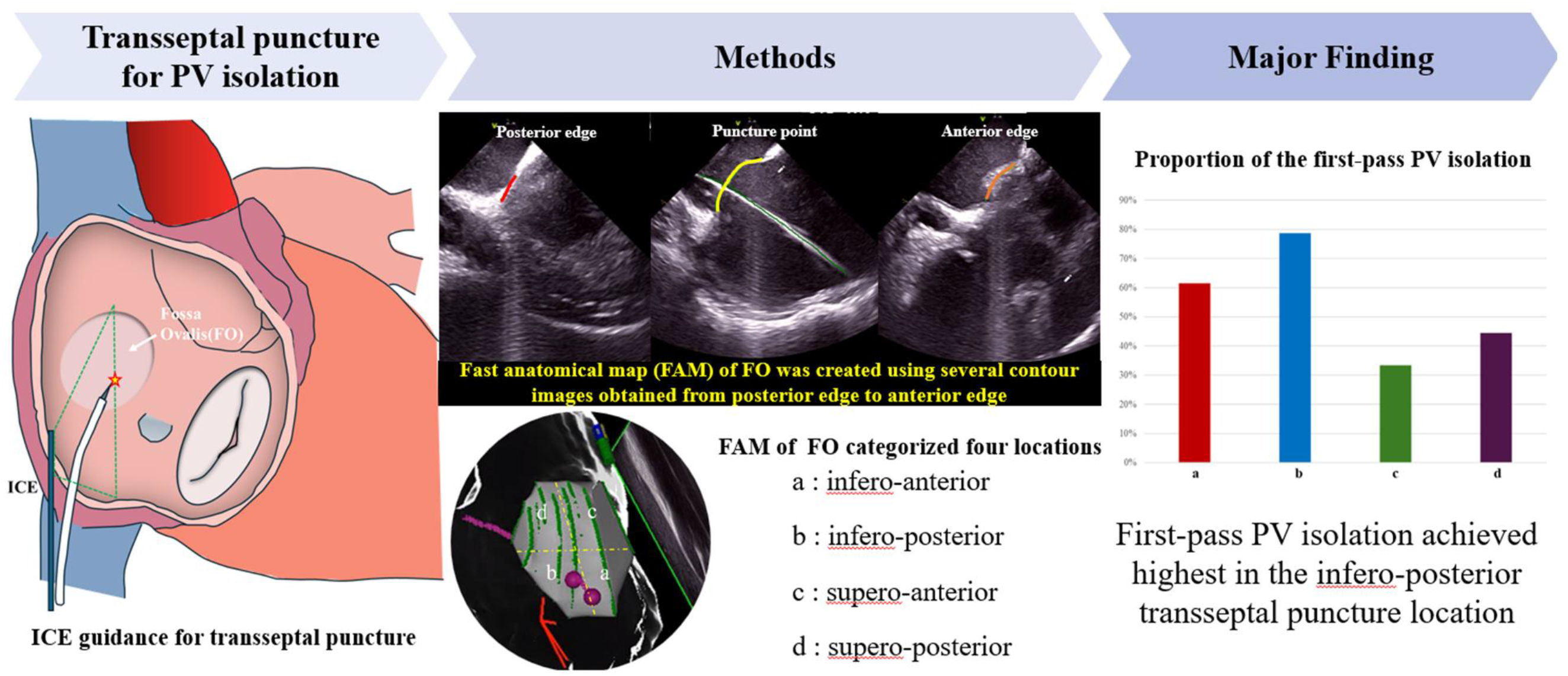

## Introduction

Atrial fibrillation (AF), which induces heart failure and systemic embolization is the most common arrhythmia detected in clinical practice.^1^ To reduce these risks, treatment such as antiarrhythmic drug therapy and catheter ablation are performed. In particular, as its higher success rate than that of antiarrhythmic drug therapy, catheter ablation has emerged as a first-line therapy over the past 10 years.^2^ Actually, catheter ablation has been shown to reduces stroke,^3, 4^ heart failure, and all-cause mortality. ^5^ ^6^

Regarding the source of AF, the pulmonary vein (PV) has been identified as the most common trigger site for AF.^7^ Therefore, PV isolation in which the PV is electrically isolated by creating circumferential lesions around the PV is clearly stated in current guideline.^8^ According to some recent studies, achieving first-pass PV isolation– PV isolation in which no residual conduction gap remains following initial circumferential lesion is created around the PV–has proven to lead to better results in terms of AF recurrence during follow-up.^9, 10^ Therefore, to avoid the creation residual conduction gap, forming sufficient lesions using radiofrequency ablation catheters have focused recently. Although the ablation index (AI)–index parameter of lesion formation which combines output, ablation duration, and contact force–,^11, 12^ interlesion distance,^13, 14^ catheter stability,^15^ and catheter contact vector direction^16^ has been reported to affect first-pass PV isolation, these are all related to lesion formation. On the other hand, some reports have shown that the transseptal puncture location on the fossa ovalis also affects PV isolation.^17–19^ However, those reports remained only suggested an ideal transseptal puncture location or determined the location using 2D image of transesophageal echocardiography images or computed tomography images which frequently pointed out fusion errors when images are merge into 3D mapping system.^20^ Thus, the optimal transseptal puncture location for successful first-pass PV isolation has not been fully assessed so far.

Currently available intracardiac echocardiography (CARTO SOUND: Biosense Webster, Irvine, CA, USA) enables not only the identification of cardiac structures, but also the representation of 3D geometry of these structures as fast anatomical map (FAM) on 3D mapping system (CARTO 3 system: Biosense Webster, Irvine, CA, USA).^21^ As a result, this technology enables the precise identification of the transseptal puncture location on fossa ovalis.

In this present study, we aimed to investigate the relationship between the transseptal puncture location on the fossa ovalis and the first-pass PV isolation success rate using this system.

## Method

### Study Population

This retrospective study population comprised consecutive 102 patients who had undergone their first catheter ablation for AF at Kumamoto University Hospital between December 2022 and October 2023. Before admission, adequate oral anticoagulation was maintained for at least 1 month before the procedure. Warfarin was not interrupted during the procedure. Dabigatran, rivaroxaban, apixaban, and edoxaban were omitted only on the morning of the procedure. The absence of a thrombus in the left atrium was confirmed through transesophageal echocardiography or contrast-enhanced computed tomography before the procedure.

This study adhered to the Declaration of Helsinki principles and received approval from the Institutional Review Board and Ethics Committee of Kumamoto University (approval number 2772). Informed consent was waived due to the low risk of the cohort study and the impracticality of obtaining direct consent from all subjects. Instead, the study protocol was extensively publicized at Kumamoto University Hospital and on the website, allowing patients to opt-out if they wished.

### Study protocol

During the procedure, transseptal puncture was performed under intracardiac echocardiography guidance using a transseptal puncture needle (RF needle: Boston Scientific, Marlborough, MA, USA) via an 8.5-Fr long sheath (Daig SL-0: Abbot, Chicago, IL, USA). The edge of the guidewire was placed in the left PV, and the contour of the guidewire was created using intracardiac echocardiography images where the puncture area was clearly confirmed. Subsequently, FAM of the fossa ovalis was created using several contour images obtained using intracardiac echocardiography from the anterior edge to the posterior edge. Figure 1 illustrate the method used to confirm the transseptal puncture location on the fossa ovalis. The enrolled patients were then categorized into four groups based on the transseptal puncture location of the fossa ovalis: infero-anterior, infero-posterior, supero-anterior, and supero-posterior (Figure 1).

**Figure 1.**
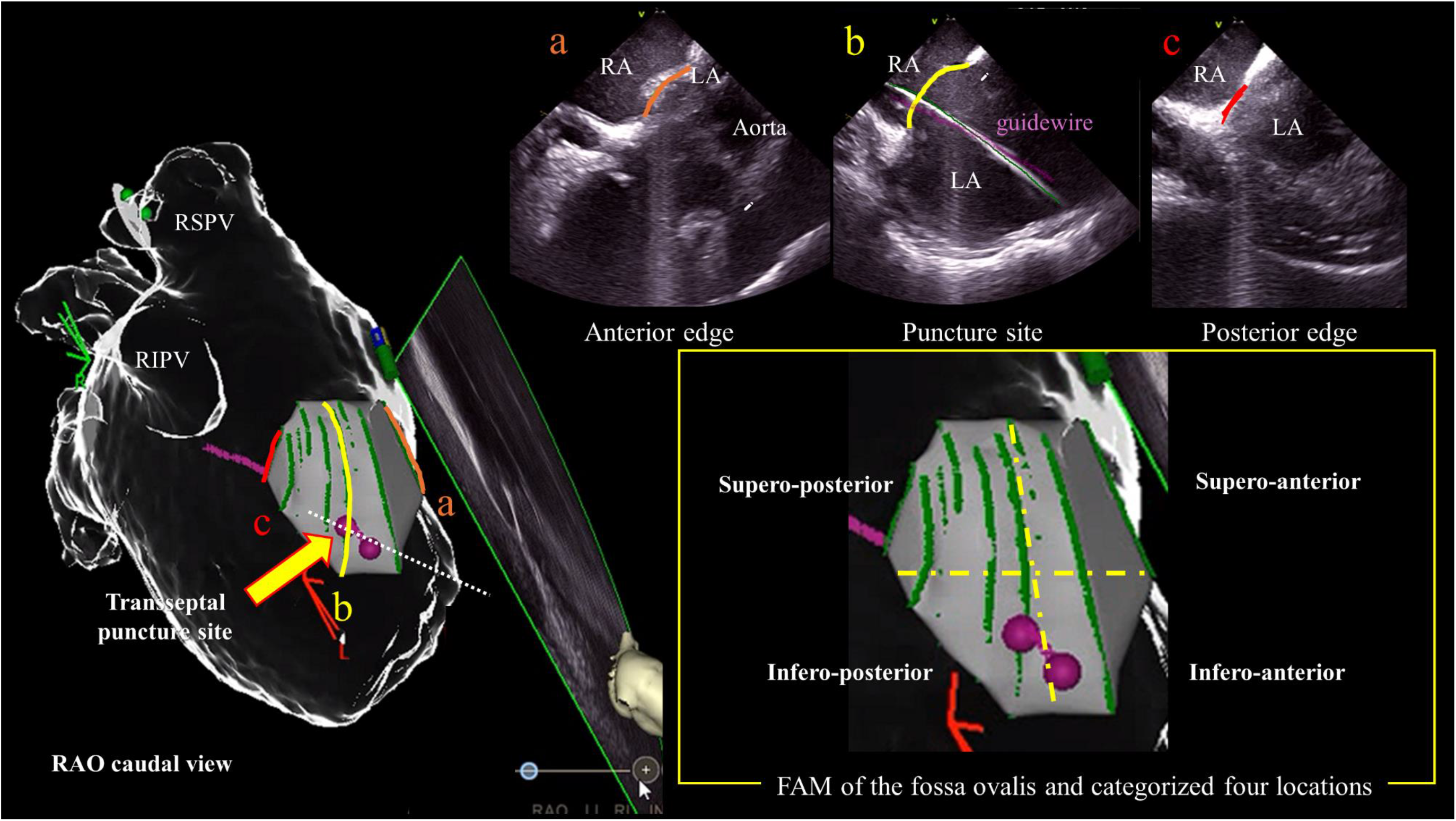
Categorization of transseptal puncture location based on fast anatomical map (FAM) of the fossa ovalis. The FAM of the fossa ovalis was created using intracardiac echocardiography images from the anterior edge (a) to posterior edge (c). The point of intersection between the guidewire and fossa ovalis (b) was defined as the transseptal puncture site, which was classified into four locations as shown.Abbreviations: LA, left atrium; RA, right atrium; RAO, right anterior oblique; RIPV, right inferior pulmonary vein; RSPV, right superior pulmonary vein.

The primary evaluation point was the comparison of the first-pass PV isolation rates among the four groups. And the secondary evaluation point was comparison of the ablation index (AI) value and proportion of the contact vector is directed proximal (P-vector)–one of the forms of catheter contact vector direction which represents insufficient catheter contact^16, 22^– among four groups. Moreover, these analysis parameters were evaluated in six segments of each PV (supero-anterior, roof, supero-posterior, infero-posterior, bottom, and infero-anterior). The target AI values were 350–400 for the posterior wall, 400–450 for the non-posterior wall under inter lesion distance was 4–5 mm following previous report. ^23, 24^

For the AI value, the mean values were compared among the four groups. And the values in the four groups were also evaluated to determine whether the value met the target AI value.

Regarding the contact vector direction, it has been shown that, if the contact vector is exhibited P-vector against distal tip of the catheter, the lesion size is significantly smaller than that of in other vectors owing to insufficient catheter tip contact.^16^ Therefore, we divided the contact vector direction into a P-vector and other vectors (normal-vector).^16, 22^ Furthermore, the segments in which the P-vector was observed were distinguished from those in which only the normal-vector was detected following methods described in previous studies.^16, 22^ The proportion of P-vector segments was compared between the four groups for each PV segment.

### Catheter ablation

Catheter ablation was performed under moderate to deep sedation using a bolus dose of morphine and fentanyl in combination with a continuous infusion of dexmedetomidine hydrochloride. Patient respiration was controlled using bi-level positive airway pressure in all cases. Intravenous heparin was administered to maintain an activated clotting time of between 300 and 350s. Before transseptal puncture, a 6-French duo-decapolar steerable catheter (BeeAT, Japan Lifeline, Tokyo, Japan) was percutaneously inserted with the tip positioned in the coronary sinus via the right jugular vein. Then, following transseptal puncture performed as described above, an 8.5-Fr long sheath (Daig SL-0; Abbot) and an 8.5-Fr steerable sheath (VIZIGO; Biosense Webster) were inserted into the left atrium. Before PV isolation, left atrial FAM was created using a multipolar catheter (PENTARAY or OCTARAY; Biosense Webster). PV isolation was performed using FAM guidance. During PV isolation, radiofrequency energy was delivered at 35–40 W using an irrigation catheter (ThermoCool SmartTouch SF catheter; Biosense Webster). The target AI values were 350–400 for the posterior wall, 400–450 for the non-posterior wall under inter lesion distance was 4–5 mm following previous report.^23, 24^ After completion of the initial PV isolation line, isolation was confirmed by the absence of PV potentials recorded on the multipolar catheter electrodes and no electrical signals from the veins entering the atrium remained (bidirectional block). If the initial ablation line did not create a conduction block, additional radiofrequency ablations was performed to complete the PV isolation.

The parameters of automated tag setting were set as follows: (1) catheter stability range of motion, < 3 mm; (2) catheter stability duration, > 3 s; (3) contact force, > 3 g, time, > 25%; (4) tag size, 4 mm in diameter.

### Statistical analysis

Continuous variables were compared using parametric and non-parametric test based on data distribution. All continuous data exhibited a skewed distribution, except for body mass index, left atrial diameter, and AI value, as confirmed by the Shapiro– Wilk test. Therefore, continuous data, except for body mass index, left atrial diameter, and AI value were expressed as medians (interquartile range) and were analyzed using non-parametric tests. Differences in these variables among multiple groups were assessed using the Kruskal–Wallis test. On the other hand, body mass index, left atrial diameter, and AI values were expressed as the mean ± standard deviation and were analyzed using parametric tests. Differences in these parameters among multiple groups were assessed using one-way analysis of variance (ANOVA). Categorical data were presented as numbers or percentages. Categorical variables were compared using Fisher’s exact test due to the small number of events. A P-value <0.05 denoted a statistically significant difference among the four groups. All statistical analyses were performed using R (R Foundation for Statistical Computing, Vienna Austria).

## Results

### Patients’ characteristic

Within this investigation, the number of transseptal puncture area located in the infero-anterior, infero-posterior, supero-anterior, and supero-posterior regions of the fossa ovalis were 26 (25.5%), 61 (59.8%), 6 (5.9%), and 9 (8.8%) patients respectively. Table 1 summarizes the patients’ characteristics. Notably, no significant differences in age, sex, body mass index, estimated glomerular filtration rate, B-type natriuretic peptide, ejection fraction, proportion of non-paroxysmal AF, hypertension proportion, diabetes mellitus proportion, CHADS_2_ score, and left atrial diameter were found among the four groups.

**Table 1.** Patient characteristics

|  | All<br>(n=102) | Infero-anterior<br>(n=26) | Infero-posterior<br>(n=61) | Supero-anterior<br>(n=6) | Supero-posterior<br>(n=9) | P-value |
| --- | --- | --- | --- | --- | --- | --- |
| Age, year | 70 (62.3–73) | 70.5 (64.3–74.8) | 69 (59–73) | 65.5 (64–71.5) | 72 (62–74) | 0.7 |
| Male sex, n (%) | 72 (70.6%) | 20 (76.9%) | 44 (72.1%) | 3 (50.0%) | 5 (55.6%) | 0.4 |
| BMI, kg/m<sup>2</sup> | 24.3±3.3 | 23.7±3.1 | 24.1±3.4 | 25.6±3.6 | 25.7±3.8 | 0.31 |
| Non-paroxysmal, n (%) | 46 (43.0%) | 13 (50.0%) | 24 (39.3%) | 3 (50.0%) | 6 (66.7%) | 0.41 |
| eGFR, mL/min/1.73 m<sup>2</sup> | 61 (50–72) | 56 (48–68.3) | 65 (52–81) | 63.5 (54–64) | 56 (42–66) | 0.21 |
| BNP, pg/mL | 62.6 (23.3–131.3) | 57.4 (24.1–149.7) | 51.1 (21.6–113.9) | 80.6 (30.8–117.2) | 123.5 (67.3–221.3) | 0.13 |
| EF, % | 60.1 (54.2–64.4) | 58.6 (45.6–63.8) | 60.3 (55.2–63.4) | 62.8 (60.5–65.1) | 61.1 (54.2–64.6) | 0.35 |
| HT, n (%) | 72 (70.6%) | 20 (76.9%) | 40 (65.6%) | 5 (83.3%) | 7 (77.8%) | 0.7 |
| DM, n (%) | 15 (14.7%) | 2 (7.7%) | 11 (18%) | 1 (16.7%) | 1 (11.1%) | 0.67 |
| History of stroke, n (%) | 9 (8.8%) | 4 (15.4%) | 4 (6.6%) | 1 (16.7%) | 0 (0%) | 0.29 |
| CHADS2 score | 1 (1–2) | 2 (1–2.8) | 1 (1–2) | 1.5 (1–2) | 1 (1–2) | 0.36 |
| LADs, mm | 39.7±6.9 | 40.6±6.5 | 38.4±6.6 | 42.9±9.4 | 43.7±6.9 | 0.08 |

- Abbreviations: BMI, body mass index; BNP, B-type natriuretic peptide; CHADS2, Congestive Heart Failure, Hypertension, Age $\geq 75$ years, Diabetes Mellitus, Stroke (doubled); DM, diabetes mellitus; EF, ejection fraction; eGFR, estimated glomerular filtration rate; HT, hypertension; LAD, left atrial diameter.

### Relationship between transseptal puncture location and residual conduction gap

All PVs were successfully isolated eventually. However, residual conduction gaps after the creation of an initial PV isolation line were identified in 32 of 102, with a total of 50 residual conduction gaps. Figure 2 represents the proportion of the first-pass PV isolation in each group. As shown in this figure, first-pass PV isolation was achieved in 16 of 26 (61.5%) patients in infero-anterior, 48 of 61 (78.7%) patients in infero-posterior, 2 of 6 (33.3%) patients in supero-anterior, and 4 of 9 (44.4%) patients in supero-posterior transseptal puncture location, respectively (P = 0.02). Notably, the infero-posterior group had the highest achievement proportion in first-pass PV isolation. Table 2 represent the locations of the residual conduction gaps in each group. As shown in this table, a residual conduction gap was frequently observed on the inferior side of the right PV in every groups.

**Figure 2.**
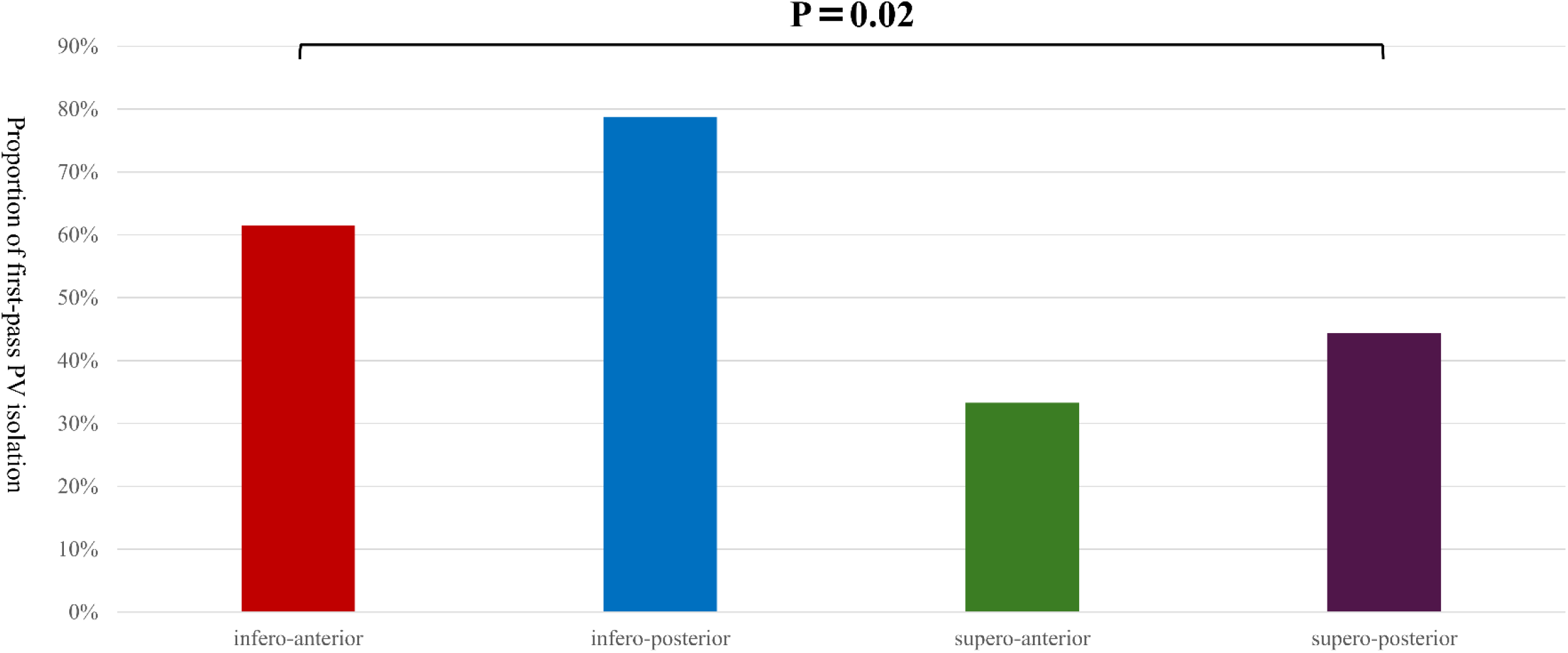
Frequency of residual conduction gap following creating the initial circumferential PV isolation line among the four groups. Abbreviation: PV, pulmonary vein.

**Table 2.**
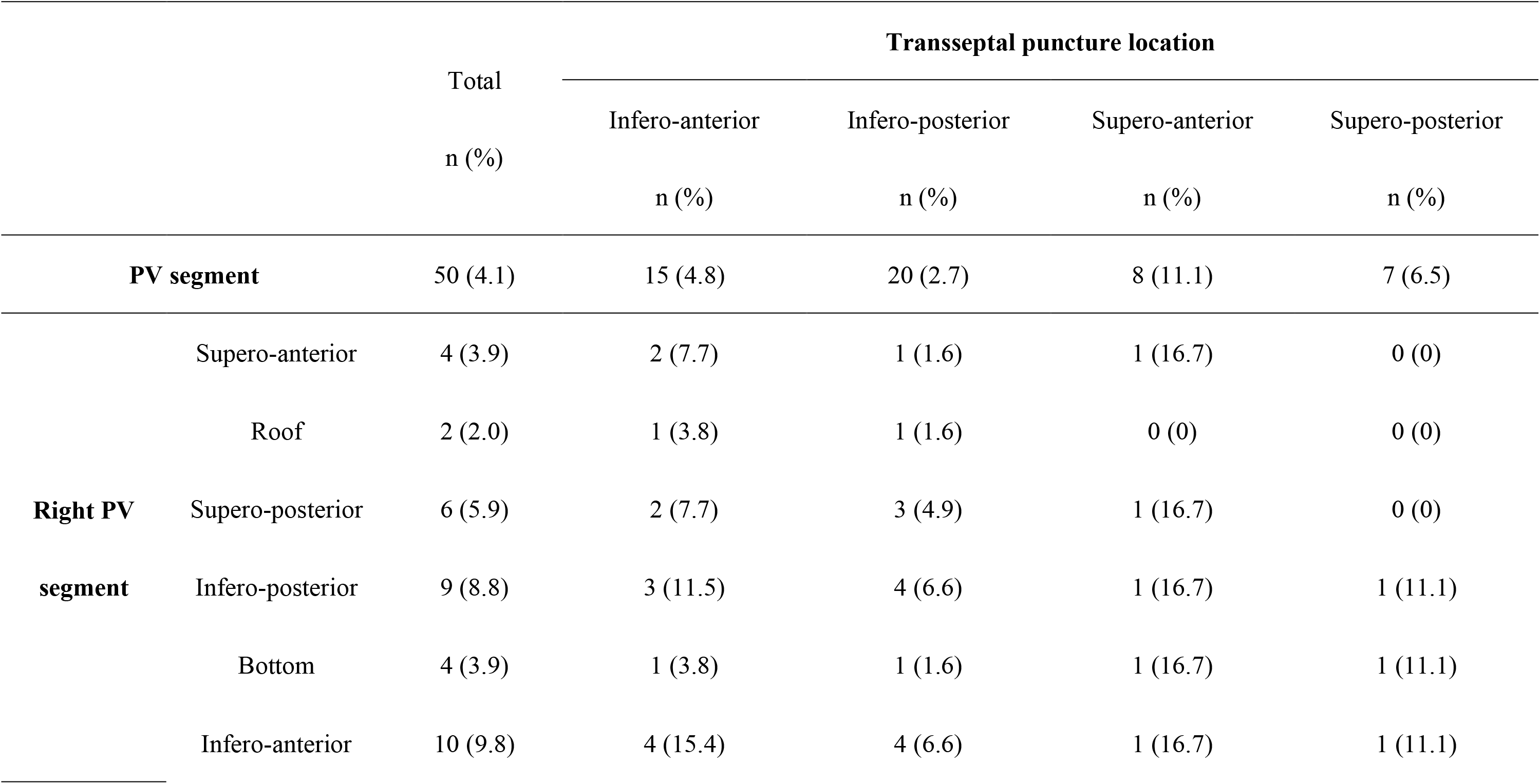
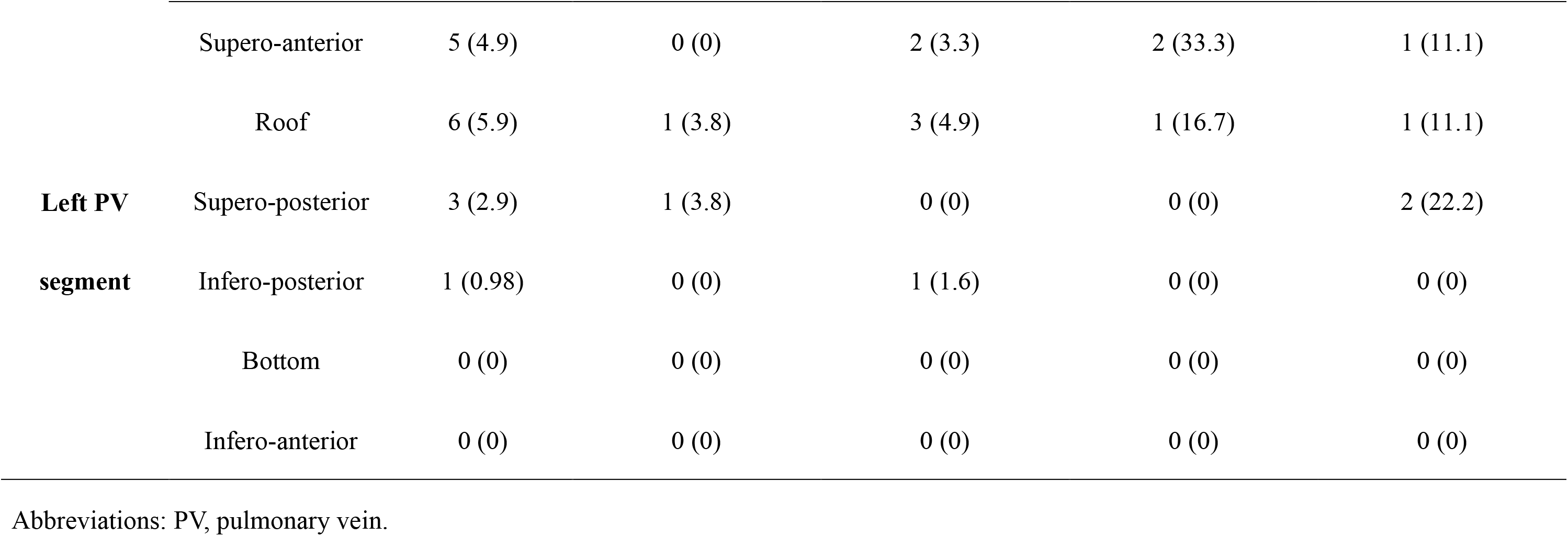
Residual conduction gaps in each segment

Procedure time for the infero-anterior, infero-posterior, supero-anterior, and supero-posterior transseptal puncture groups were 125 (119–134), 119 (103–136), 142 (129–152), and 144 (126–150) min, respectively (P = 0.048). The procedure time was the shortest in the infero-posterior group.

### Relationship between transseptal puncture location and AI value

Figure 3 summarizes the mean patient’ AI values in each group. The mean AI value did not significantly differ among the four groups (infero-anterior 401.6±7.3 vs. infero-posterior 401.9±5.2 vs. supero-anterior 397.5±4.7 vs. supero-posterior 398.6±5.3, P = 0.17). The mean AI values for each PV segment in the four groups are provided in Table 3. No significant differences among the groups were seen for each PV segments, except for the right PV supero-posterior segment (P = 0.03), right PV infero-posterior segment (P = 0.01), left PV supero-anterior segment (P = 0.03), and left PV bottom segment (P = 0.03). In these segments, the upper puncture groups (supero-anterior and supero-posterior) had a lower AI than the lower puncture groups (infero-anterior and infero-posterior). However, except for the left PV bottom segment in the supero-anterior puncture group, the AI value of each segment met the target AI value.

**Figure 3.**
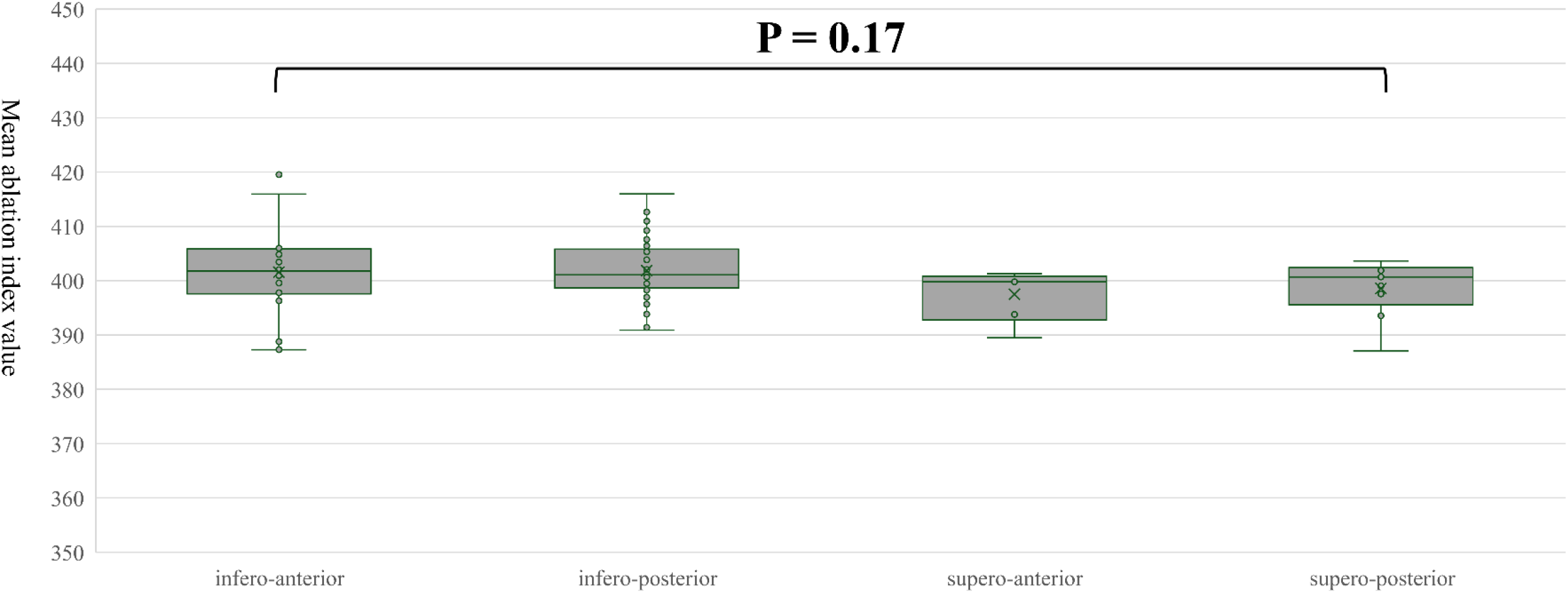
Mean AI value among four groups. Abbreviation: AI, ablation index.

**Table 3.**
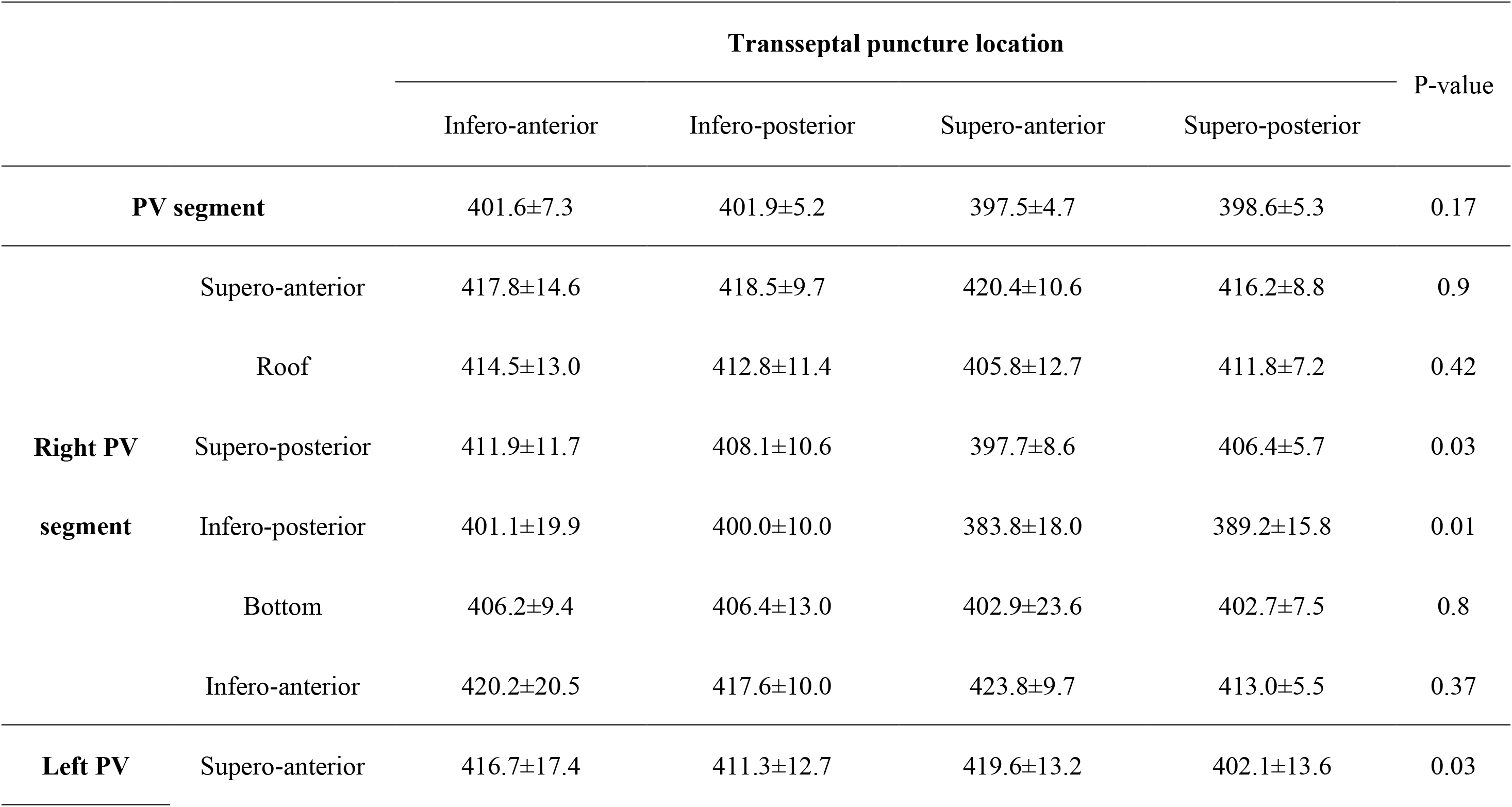
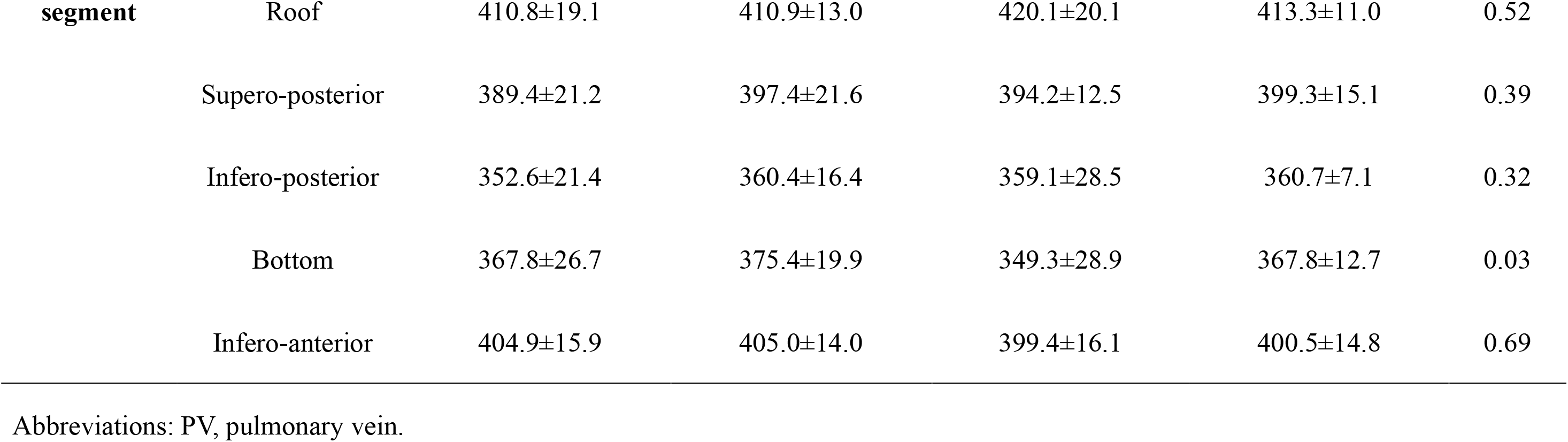
Mean AI values in each segment among the four groups

### Relationship between transseptal puncture location and P-vector segments

Among all 1,224 PV segments in 102 patients, the P-vector was observed in 134 segments (10.9%). In these 134 segments, 17 (12.7%) residual conduction gaps were detected. In comparison, in the remaining 1,090 segments in which only the normal-vector was observed, 33 (3.0%) residual conduction gaps were noted. The residual conduction gap was significantly more frequently discerned in segments in which the P-vector was observed than in segments in which only the normal-vector was observed (P < 0.001). This finding was consistent with the previous report related to the contact vector direction; the P-vector indicated insufficient contact against target tissue.^16^

Figure 4 shows the proportion of segments in which the P-vector was observed in each group. The number and proportion of the segment in which the P-vector was observed in the infero-anterior, infero-posterior, supero-anterior, and supero-posterior transseptal puncture groups were 48 (15.4%), 63 (8.6%), 12 (16.7%), and 11 (10.2%) segments, respectively (P < 0.01). As shown in Figure 4, the infero-posterior transseptal puncture group had the lowest proportion of P-vector was observed.

**Figure 4.**
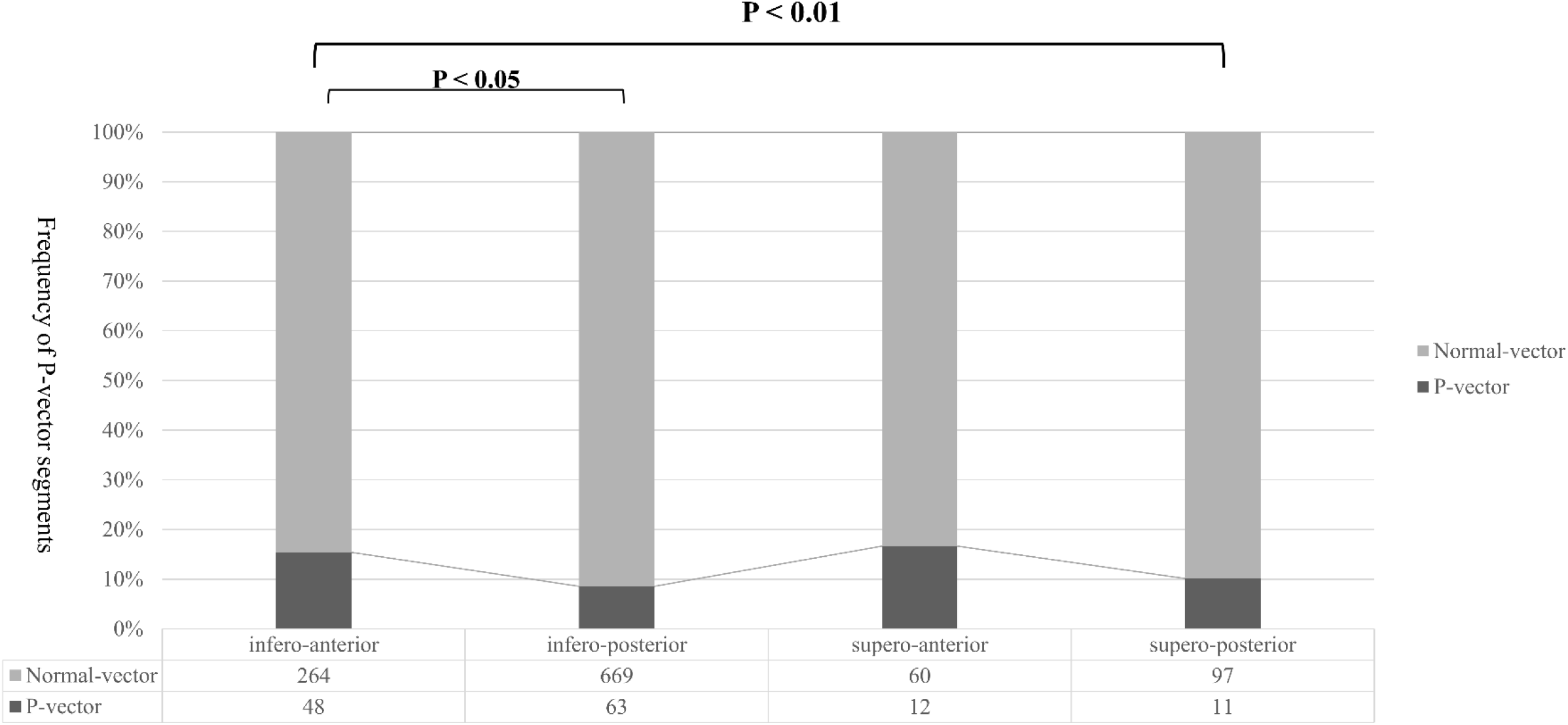
Frequency of P-vector segment among four groups. Abbreviation: P-vector, proximal catheter contact vector.

Table 4 represents the proportion of P-vectors in each segment among four the groups. As shown in this table, no significant difference was seen among the groups for each PV segment, except for the infero-anterior segment of right PV (P < 0.05). Notably, this segment exhibited a higher tendency for formation of a residual conduction gap (Table 2). Regarding the association between the P-vector segment and the residual conduction gap in each PV segment, a significant difference was observed only at the infero-anterior segment of the right PV (P < 0.001).

**Table 4.**
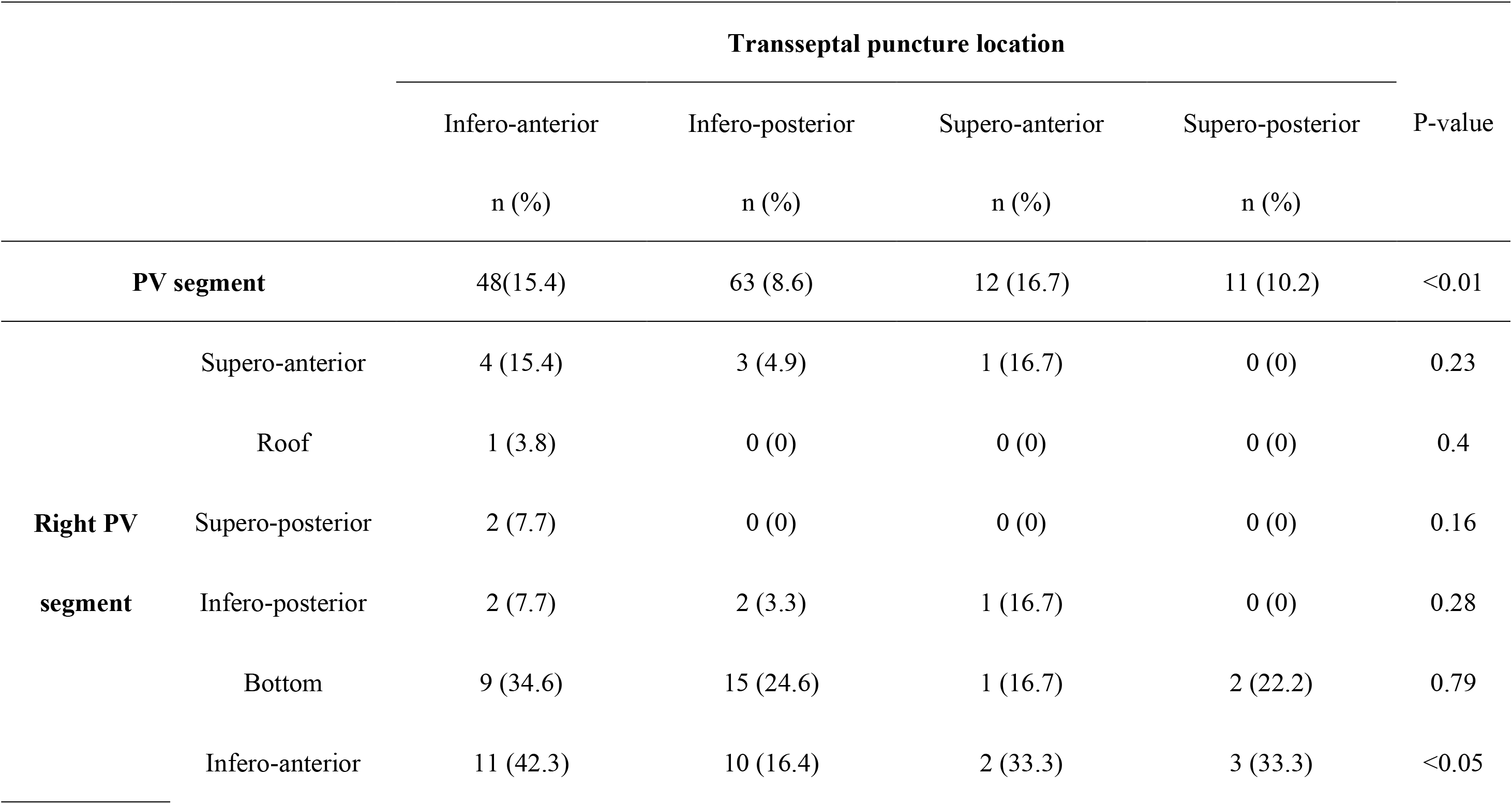
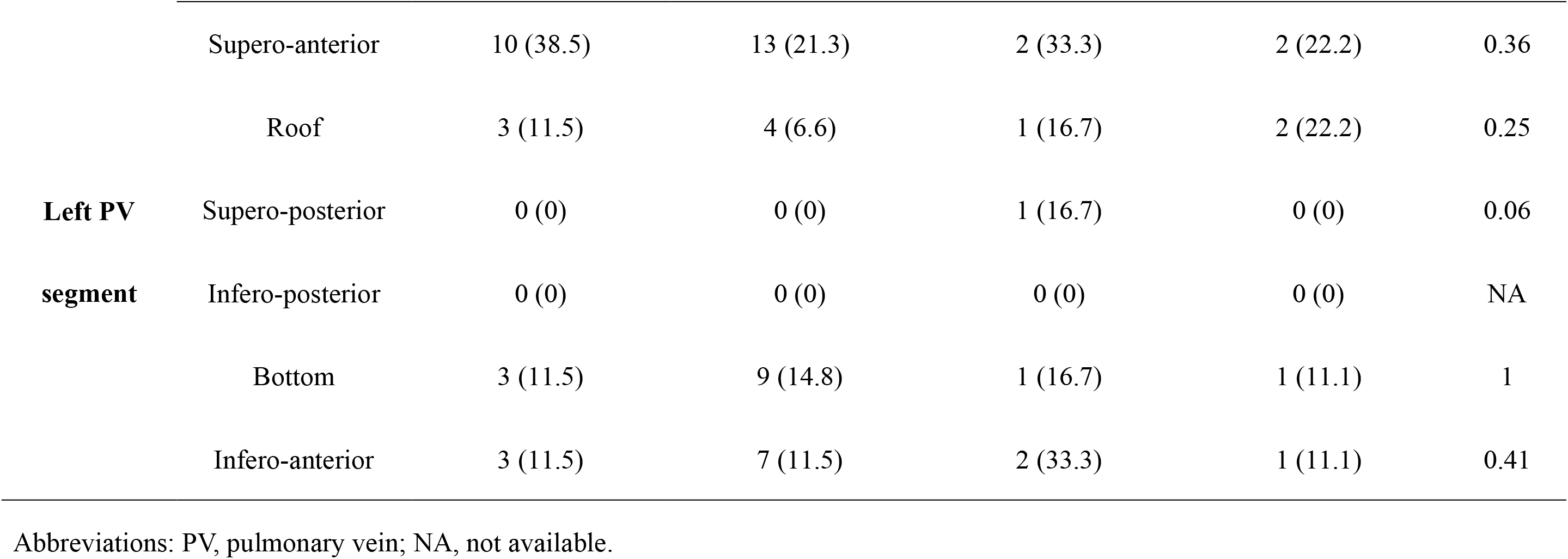
P-vector segments in each segment among the four groups

## Discussion

### Major Findings

The present study yielded the following two major results. First, successful first-pass PV isolation varied significantly depending on the transseptal puncture location on the fossa ovalis. Second, although almost all PV segments met the target AI value, and AI values did not differ significantly among four groups, the frequency of the P-vector was varied significantly depending on the transseptal puncture location.

Moreover, the infero-posterior puncture group which observed the highest first-pass PV isolation proportion, also showed the lowest proportion of the presence of a P-vector segment.

### Importance of first-pass PV isolation

Recently, although the technology related to catheter ablation has improved, AF recurrence following radiofrequency catheter ablation has been reported to be high in approximately 10–20% of the patients.^25^ Some reports have suggested that this is owing to non-PV foci, atrial substrates, or lifestyle factors.^26^ However, PV electrically reconnection has also been shown to contribute to recurrence, even in the current era.^27^ This PV electrically reconnection has reported to close relationship to failure of first-pass PV isolation. Consequently, first-pass PV isolation is considered to be an important factor in achieving durable PV isolation and has been the focus of recent research^28^. To achieve first-pass isolation, target AI value, and catheter stability have been discussed.^11, 12, 15^ However, even when these factors have been considered, the first-pass PV isolation rate has varied between 69 to 88%.^9, 23^ This finding was consistent with the result of the present study. Therefore, other unrecognized factors related to first-pass PV isolation may be exist. In this present study, the transseptal puncture location on the fossa ovalis significantly affected first-pass PV isolation. This finding suggests that the location of the transseptal puncture on the fossa ovalis may have influenced first-pass PV isolation, even after considering the AI value and catheter stability during ablation.

### Relationship between AI value and transseptal puncture location

In this present study, although the mean AI value at almost all segments met the target AI value, the value of the right PV posterior segments and left PV supero-anterior and bottom segments among superior puncture location groups were smaller than those of the inferior puncture location groups. This phenomenon may be attributed to insufficient contact. Actually, although the operators aimed to achieve higher AI at those segments, complete ablation failed in the superior puncture location groups, owing to catheter movement. Moreover, the residual conduction gap in these segments tended to be higher in these puncture location groups. Therefore, when transseptal puncture is performed in the superior locations, the operator’s intended AI value may not be achieved leading to residual conduction gap forming

### Relationship between P-vector and transseptal puncture location

It has been shown that, although sufficient contact force is detected, only the proximal edge, but not the distal tip of the catheter touches the tissue in cases where the catheter contact vector indicates the P-vector. This contact vector direction has been found to be closely related to forming residual conduction gap.^16, 29^ Therefore, once the P-vector is observed during ablation, it is important to pull the catheter back until changes to a normal-vector. Although the operators recognizing this phenomenon, the P-vector was observed at a certain rate in this present study. The proportion of P-vectors differed significantly depending on the transseptal puncture location. The proportion of the P-vectors was significantly lower in the infero-posterior puncture location group than in the other puncture groups. This phenomenon was clearly observed particularly in the infero-anterior segment of the right PV. In this segment, the length from the transseptal puncture location to this segment was the closest among the various segments. Consequently, adjustment of the catheter contact situation might be difficult, leading to ablation in P-vector situation even when the contact vector direction is recognized.

### The optimal transseptal puncture location for PV isolation

The previous studies have discussed the relationship between the transseptal puncture location and PV isolation success rate. However, one of these report only suggested an ideal transseptal puncture location,^30^ while the other reported that PV isolation success rate was not dependent on the transseptal puncture locations (anterior, medial, or posterior) determined using transesophageal echocardiography. Although the anatomy of the fossa ovalis is structured in 3D, that investigation evaluated fossa ovalis only in 2D.^17^ Therefore, the relationship between the transseptal puncture location and PV isolation success rate had not been fully evaluated so far. In this present study, this relationship was evaluated using the 3D structure of the fossa ovalis. The data showed that the infero-posterior puncture location was the most effective transseptal puncture location for PV isolation in radiofrequency catheter ablation due to the lower residual conduction gap proportion than in the other groups. The procedure time of the infero-posterior puncture location group was lower than that of the other groups in this present study (P = 0.048). This may be due to a reduction in residual conduction gaps.

Therefore, it might be important to consider the transseptal puncture location of the fossa ovalis to achieve the first-pass PV isolation, given that this location may decrease AF recurrence in the future.

### Study limitations

In this present study, fewer cases were observed in the upper location puncture groups than in the lower location puncture groups. If more cases had been included in the upper location transseptal puncture groups, different results may have been obtained.

## Conclusion

The proportion of first-pass PV isolation achievement was significantly higher in the infero-posterior puncture group. This may have been owing to the sufficient catheter contact situation than the other location groups. Therefore, the transseptal puncture location on the fossa ovalis may be an important factor in achieving first-pass PV isolation. Our results indicated that the transseptal puncture location during radiofrequency ablation for AF might affect the procedure time and AF recurrence.

## Data Availability

The datasets used and analyzed during the current study available from the corresponding author on reasonable request.

## Acknowledgments

The author would like to thank Mr. Shogo Tsuda, Mr. Kazunobu Sugama and Ms. Asahi Tominaga for technical support.

## Sources of Funding

This investigation was supported by the Johnson & Johnson Medical Research Grant and Takeda Science Foundation.

## Disclosures

Dr Tsujita has received honoraria from AMI Co., Ltd., Bayer Yakuhin, Ltd., Bristol-Myers K.K., EA Pharma Co.,Ltd., Mochida Pharmaceutical Co., Ltd., and scholarship fund from AMI Co., Ltd., Bayer Yakuhin, Ltd., Boehringer Ingelheim Japan, Chugai Pharmaceutical Co, Ltd., Daiichi Sankyo Co., Ltd., Edwards Lifesciences Corporation, Johnson & Johnson K.K., Ono Pharmaceutical Co., Ltd., Otsuka Pharmaceutical Co.,Ltd., Takeda Pharmaceutical Co., Ltd., and honoraria from Amgen K.K., Bayer Yakuhin, Ltd., Daiichi Sankyo Co., Ltd., Kowa Pharmaceutical Co. Ltd., Novartis Pharma K.K., Otsuka Pharmaceutical Co.,Ltd., Pfizer Japan Inc., and belongs to the endowed departments donated by Abbott Japan Co., Ltd., Boston Scientific Japan K.K., Fides-one, Inc., GM Medical Co., Ltd., ITI Co.,Ltd., Kaneka Medix Co., Ltd., Nipro Corporation, Terumo Co, Ltd., Abbott Medical Co., Ltd., Cardinal Health Japan, Fukuda Denshi Co., Ltd., Japan Lifeline Co.,Ltd., Medical Appliance Co., Ltd., Medtoronic Japan Co., Ltd.

## Notes

### Clinical Trial

This study is retrospective study. Therefore, no clinical trial ID was assigned.

### Author Declarations

This study adhered to the Declaration of Helsinki principles and received approval from the Institutional Review Board and Ethics Committee of Kumamoto University (approval number 2772).

